# Assessing Post-TAVR Cardiac Conduction Abnormalities Risk Using a Digital Twin of a Beating Heart

**DOI:** 10.1101/2024.03.28.24305028

**Authors:** Symon Reza, Brandon Kovarovic, Danny Bluestein

## Abstract

Transcatheter aortic valve replacement (TAVR) has rapidly displaced surgical aortic valve replacement (SAVR). However, certain post-TAVR complications persist, with cardiac conduction abnormalities (CCA) being one of the major ones. The elevated pressure exerted by the TAVR stent onto the conduction fibers situated between the aortic annulus and the His bundle, in proximity to the atrioventricular (AV) node, may disrupt the cardiac conduction leading to the emergence of CCA. In his study, an *in-silico* framework was developed to assess the CCA risk, incorporating the effect of a dynamic beating heart and pre-procedural parameters such as implantation depth and preexisting cardiac asynchrony in the new onset of post-TAVR CCA. A self-expandable TAVR device deployment was simulated inside an electro-mechanically coupled beating heart model in five patient scenarios, including three implantation depths, and two preexisting cardiac asynchronies: (i) a right bundle branch block (RBBB) and (ii) a left bundle branch block (LBBB). Subsequently, several biomechanical parameters were analyzed to assess the post-TAVR CCA risk. The results manifested a lower cumulative contact pressure on the conduction fibers following TAVR for aortic deployment (0.018 MPa) compared to baseline (0.29 MPa) and ventricular deployment (0.52 MPa). Notably, the preexisting RBBB demonstrated a higher cumulative contact pressure (0.34 MPa) compared to the baseline and preexisting LBBB (0.25 MPa). Deeper implantation and preexisting RBBB cause higher stresses and contact pressure on the conduction fibers leading to an increased risk of post-TAVR CCA. Conversely, implantation above the MS landmark and preexisting LBBB reduces the risk.

## 1. Introduction

The minimally invasive TAVR holds great promise to replace surgical aortic valve replacement (SAVR) as the gold standard for treating aortic valve stenosis. However, TAVR is still associated with certain post-procedural complications such as paravalvular leakage, thrombosis, cardiac conduction abnormalities (CCAs), and structural valve degeneration that hampers the potential of TAVR to safely extend its use to younger and lower risk patients. Among these post-procedural complications, CCA remains notably persistent, frequently requiring the implantation of a permanent pacemaker (PPI) (Alperi et al. 2021). Recent studies have indicated that PPI is associated with increased mortality rate and heart failure hospitalization (Faroux et al. 2020; Jørgensen et al. 2019). Predicting the risk of CCA therefore holds the potential to aid the clinicians in refining their preprocedural planning strategies leading to better clinical outcomes for TAVR patients.

The interplay between the electrical and biomechanical properties of the beating heart is vital for its normal functioning. Modifications to the structural dynamics, such as changes in cardiac preload and afterload, can impact the sinus rhythm through stretching of the sino-atrial node, contractility through stretching of the atrial and ventricular myocardium (the Frank-Starling effect), and the distribution of the action potential (Quinn and Kohl 2016). Experimental studies involving the expansion of chamber lumen, both in-situ and in isolated intact hearts, have provided evidence of arrhythmias induced by mechanical factors in different species including rats (Kim, White, and Saint 2012), rabbit (Franz et al. 1992), sheep (Chen et al. 2004), and humans (Levine et al. 1988). There is compelling evidence indicating that stretching on the myocardium can act as a significant contributor to mechanically induced arrhythmias. Purkinje fibers (PFs), located in the endocardial surface of the ventricles are also stretched during lumen expansion (Canale et al. 1983). Consequently, the mechanical activation of PFs presents another plausible factor in the development of mechanically induced arrhythmias, in addition to the activation of the myocardium (Ferrier 1976; Reynolds, Chiz, and Tanikella 1975; Thakur et al. 1996). A recent study has demonstrated evidence supporting the role of PFs in conduction delay (increased R-R interval) in the hearts of rats and sheep, using targeted PF experiments (Hurley et al. 2023). In the human heart, the His bundle, positioned below the membranous septum, between the right and non-coronary cusp, may interact with the TAVR device stent. Considering the location of the TAVR anchorage zone and the nature of the conduction abnormalities observed post-TAVR, it is believed that these abnormalities are caused by the mechanical stresses exerted on the His bundle.

Several clinical studies have been conducted to identify the risk factors and improve pre-procedure planning of TAVR by determining the likelihood of post-TAVR conduction abnormalities (CCA) (Boonyakiatwattana et al. 2022; Breitbart et al. 2021; Hamdan et al. 2015; Mauri et al. 2016; Fujita et al. 2016; Maeno et al. 2017; Pollari et al. 2019; Schaefer et al. 2018; Oestreich et al. 2018; Tretter et al. 2019; Miyashita et al. 2023; Ravaux et al. 2021; Zheng et al. 2023). Clinical studies have revealed associations between post-TAVR CCA and preexisting right bundle branch block (RBBB) (Fujita et al. 2016; Hamdan et al. 2015; Maeno et al. 2017; Mauri et al. 2016; Oestreich et al. 2018; Pollari et al. 2019; Schaefer et al. 2018), deeper implantation (Fujita et al. 2016; Hamdan et al. 2015; Maeno et al. 2017; Mauri et al. 2016; Oestreich et al. 2018), shorter membranous septum length and lower ΔMSID (subtraction of implantation depth from the length of membranous septum) (Hamdan et al. 2015; Maeno et al. 2017; Oestreich et al. 2018). However, due to complex anatomical features in the device landing zone, accurately assessing the risk of CCA solely based on anatomical and procedural parameters is challenging, as post-TAVR CCA depends on the local dynamic interaction between the TAVR prosthesis and conduction fibers. Therefore, additional patient-specific computational modeling may provide more accurate predictions of patient specific CCA risk. Recently, computational studies on patient-specific structural models have indicated that elevated contact pressure and contact pressure index (CPI) on the His bundle region in the intraventricular septum below the triangular space between the RCL and NCL for self-expandable TAVR cases (Rocatello et al. 2018) and high area-weighted average logarithmic strain (AMPLS) and Contact force for balloon expandable TAVR cases were associated with onset of CCA following TAVR procedure (Reza et al. 2022). To gain more insight into the mechanisms underlying post-TAVR CCA development and provide a more accurate assessment of CCA risk, it is essential to analyze the interaction between the TAVR prosthesis and conduction fibers within a realistic, electromechanically coupled, beating heart condition. Moreover, incorporating preexisting electromechanical asynchronies into electromechanically coupled beating heart simulations will enable a comprehensive evaluation of CCA risk. However, current simulations do not consider the impact of the structural dynamics of the beating heart and pre-existing cardiac asynchrony, which is a significant contributor to new onset of CCA after TAVR.

Among post-TAVR CCAs, new onset of perioperative left bundle branch block (NOP-LBBB) associated with self-expandable TAVR devices is the major concern (13-37%) (Regueiro et al. 2016). Some TAVR patients developing perioperative NOP-LBBB following TAVR may recover through cardiac remodeling and may not require PPI, whereas some patients develop delayed high degree atrioventricular (AV) block (Maille et al. 2022) which makes it challenging for clinicians to decide whether PPI is required for the patient or not and often leads to unnecessary PPI or prolonged hospital stay. The existing computational techniques that use static patient models can only predict the risk or perioperative risk of CCA. An electromechanically coupled computational modeling technique, on the other hand, can potentially play an important role in assessing perioperative and post-operative risk of developing new conduction disturbances following TAVR; hence, avoid unnecessary PPI and at the same time, keep the patient safe from further CCA-related complications.

A dynamic high-fidelity Multiphysics model of a four-chamber adult male human heart (**Figure 1**) (Baillargeon et al. 2014), developed under the SIMULIA Living Heart Project (Dassault Systèmes, SIMULIA Corp.) has previously been used to study various cardiovascular diseases (Wisneski et al. 2020; St Pierre, Peirlinck, and Kuhl 2022) including heart failure (Genet et al. 2016; Costabal, Choy, et al. 2019), arrhythmia (Sahli Costabal, Yao, and Kuhl 2018), effects of drug on treating these diseases (Costabal, Matsuno, et al. 2019), and performance analysis of several cardiovascular procedures including TAVR (Ghosh et al. 2020b), mitral valve repair (Galili, White Zeira, and Marom 2022; Heidari et al. 2022; Pasta et al. 2022), and left ventricular assistance device (Pasta et al. 2022). In this study, this digital twin of the human heart was used to explore the interaction between the TAVR prosthesis and the conduction fibers both during the TAVR procedure and across three cardiac cycles subsequent to the procedure, in order to gain deeper insights into the mechanisms underlying the development of new conduction abnormalities at varying implantation depths and in the presence of pre-existing CCAs. This investigation holds the potential to offer valuable insights into predicting the likelihood of persistent CCA and the potential necessity for PPI due to complete AV block.

**Figure 1:**
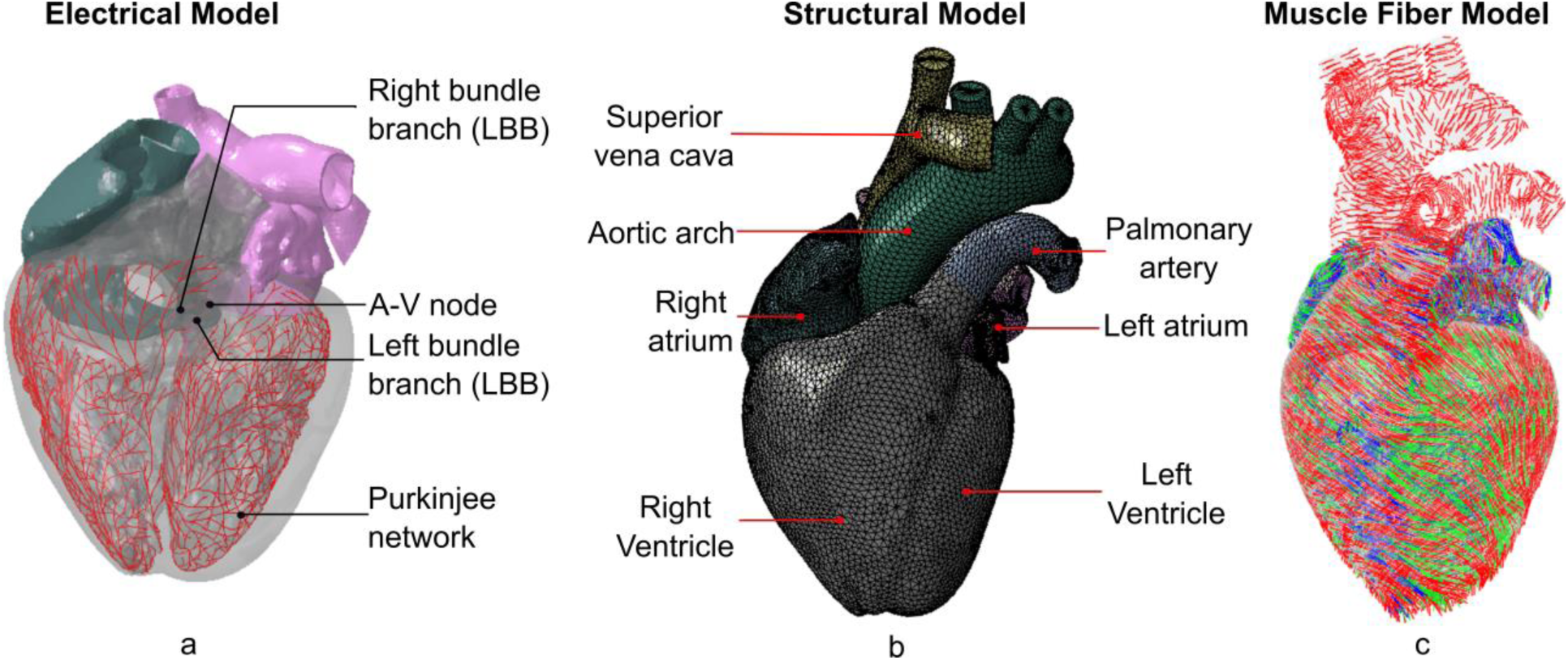
Illustration of the (a) electrical model, (b) finite element model, and (c) muscle fiber model of the 4-chamber full heart model (Adopted from SIMULIA Living Heart Human Model User Guide 2018)

## 2. Methods

A dynamic high-fidelity Multiphysics model of a four-chamber adult male human heart (Baillargeon et al. 2014), developed under the SIMULIA Living Heart Project was used as the base model for this study. This model uses a sequentially coupled electro-mechanical computational technique to simulate realistic cardiac cycles. In this study, we performed electrophysiological simulations of three different electrophysiological scenarios including healthy cardiac conduction, RBBB, and LBBB (**Figure 2**). Results obtained from the electrophysiological simulations were then coupled with the structural heart model to simulate the mechanical behavior (**Figure 3**). Simultaneously, a Evolut® 26 mm TAVR device (Medtronic, Inc., Minneapolis, MN) was deployed (**Figure 4**a-g) in a dynamic heart to assess the risk of cardiac conduction abnormality following TAVR. The recommended implantation depth (ID) for Evolut® 26 mm device is 3-6 mm. In this study, 6 mm implantation depth and normal cardiac conduction scenario were considered as the baseline. Additionally, two more implantation depths were investigated: one at 4 mm (2 mm towards the aortic side from baseline) and another at 8 mm (2 mm towards the ventricular side from baseline) in order to comprehend the effect of implantation depth in developing new conduction disturbance (**Figure 4**h). The MS length of the living heart model was 5.5 mm, yielding −1.5, +0.5, and +2.5 ΔMSID for the aortic, baseline, and ventricular implantation, respectively.

**Figure 2:**
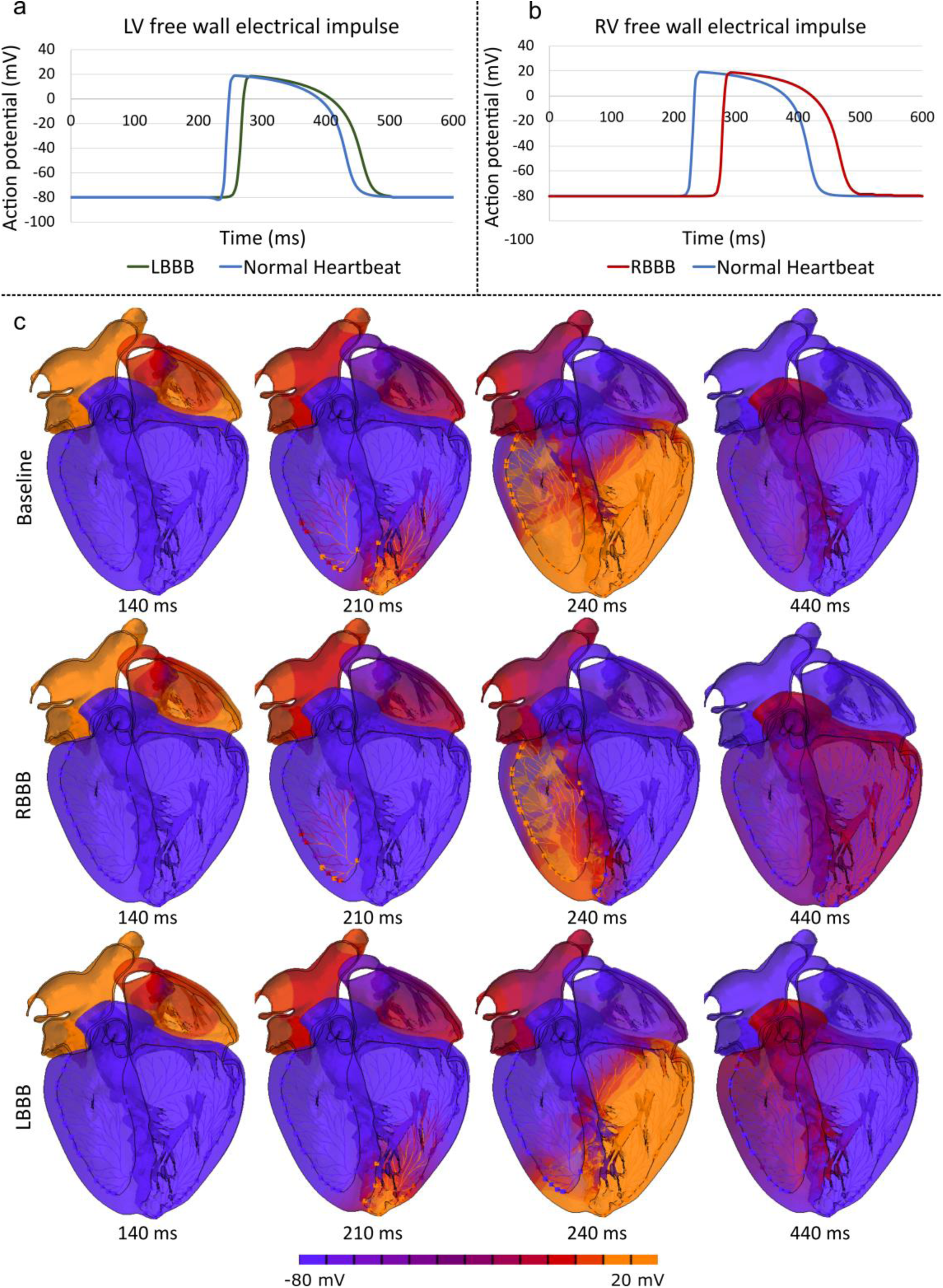
Conduction delay on the (a) left ventricular free wall due to LBBB, and (b) left ventricular free wall due to RBBB. (c) Distribution of electrical impulse at four timesteps during electrical depolarization for baseline (top row), RBBB (middle row), and LBBB (bottom row) cases.

**Figure 3:**
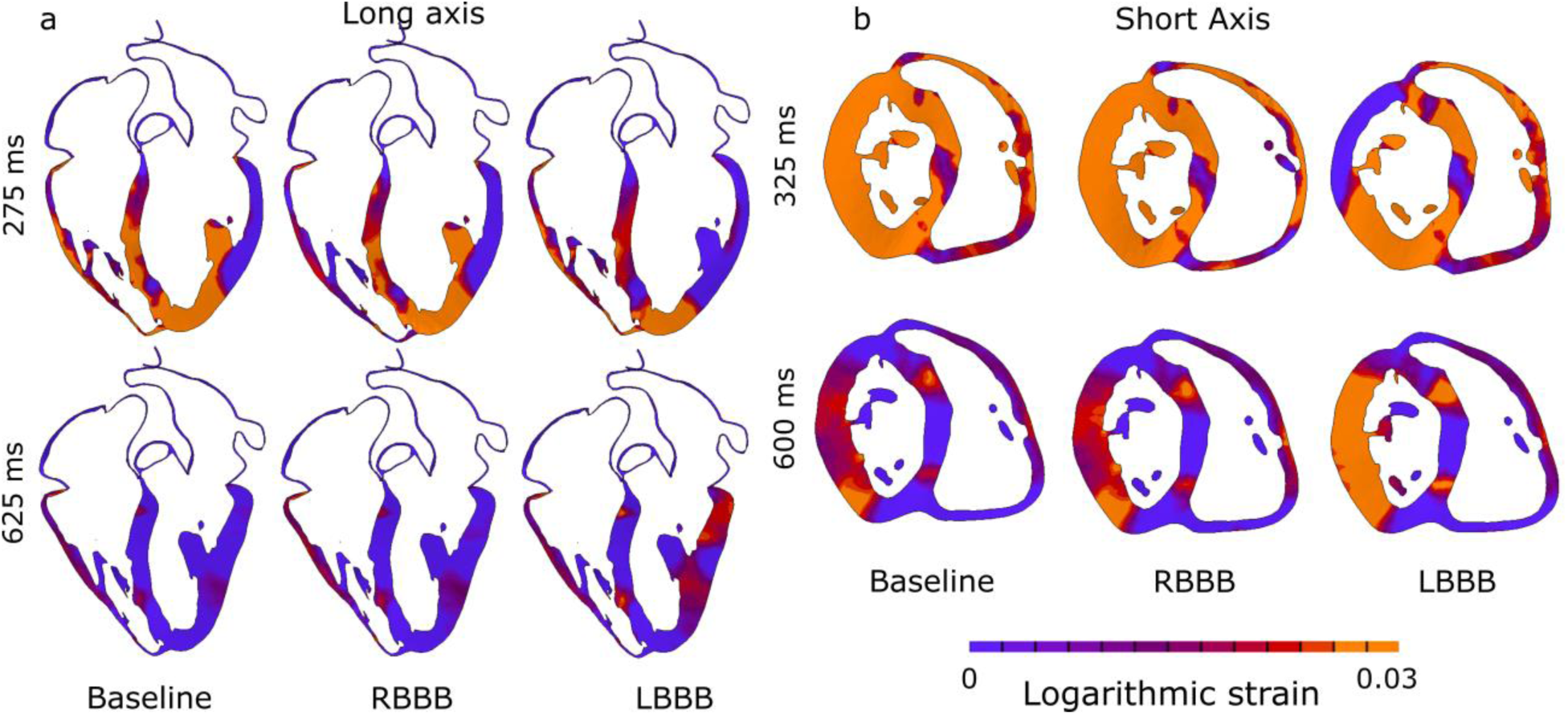
Logarithmic strain distribution for baseline, LBBB, and RBBB cases at 4 different time steps from long axis (left) and short axis (right) views.

**Figure 4:**
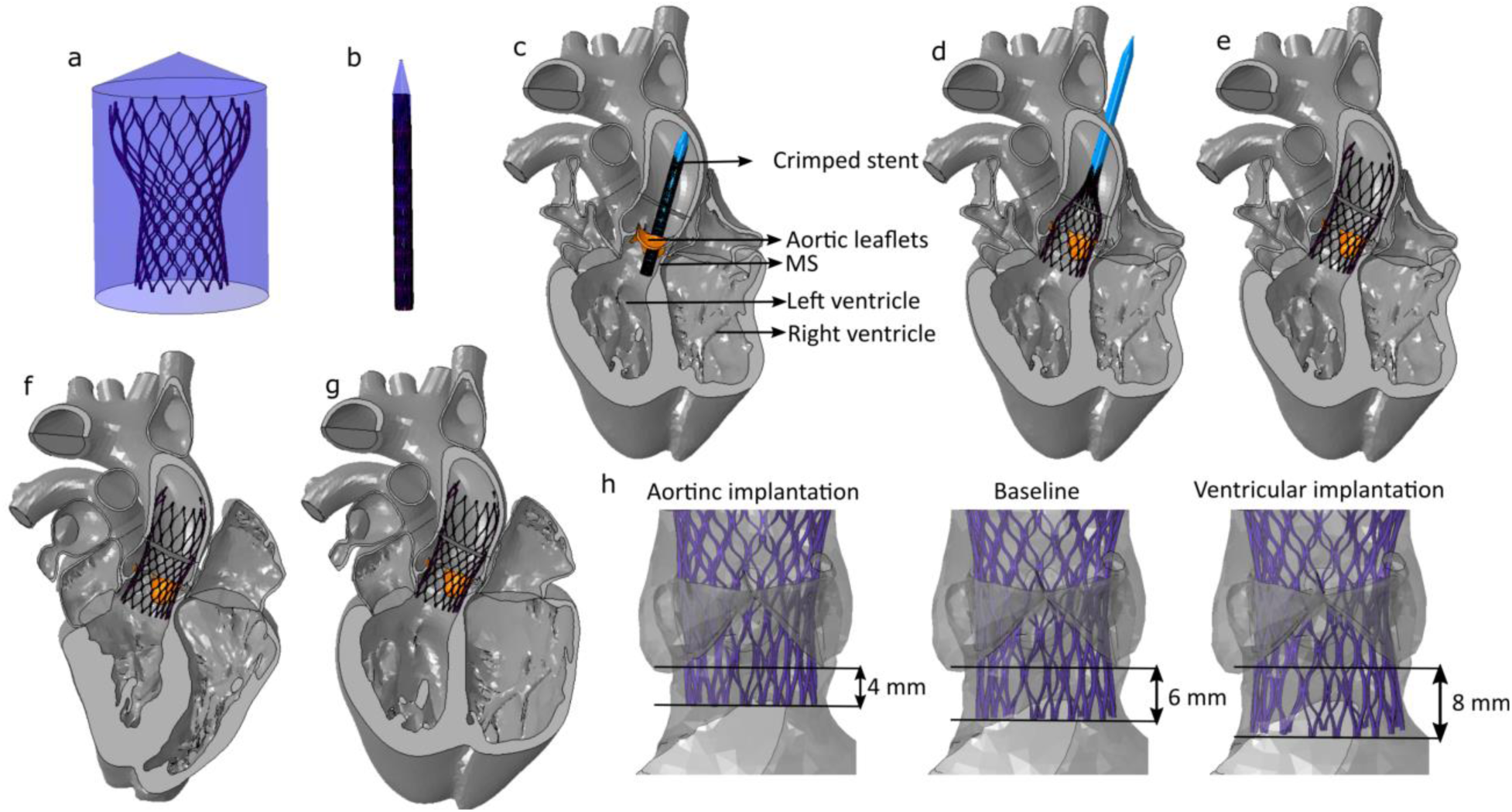
Illustration of the (a) uncrimped TAVR, (b) crimping, (c, d) implantation, (e) preload, (f) cardiac beat, and (g) relaxation steps. (h) Three considered implantation depths are also visualized.

### 2.1. Modeling cardiac electrophysiology

The Heat Transfer procedure in Abaqus/Standard is used to mimic cardiac conduction of the Living Heart Model, as the equations for diffusion heat transfer can serve as a proxy to electrical conduction equations. The electrical analysis in the cardiac cycle begins at the point of 70% ventricular diastole, prior to the atrial systole, and runs for 500 ms from the onset of depolarization to the end of repolarization across the entire heart. The temperature representing electrical potential is acquired from the simulation result which is used as a boundary condition to excite the tissue in the mechanical analysis. A bidomain model is utilized to simulate the conduction of electrical impulses through the heart, where the action potential travels through the network of Purkinje fibers and the cardiac muscles. An electrical impulse originates at the sinoatrial node (SA node) and spreads through the atrial tissue, causing atrial contraction. Sequentially, the electrical impulse is generated at the atrioventricular node (AV node) and is transmitted to the ventricles of the heart through the bundle of His. A representative geometry for the bundle of His and Purkinje fibers (Costabal, Hurtado, and Kuhl 2016) was used to conduct impulses more rapidly compared to the surrounding heart tissue. This structure is modeled as 1-dimensional electrical elements (DC1D2) with uniform conductivity, as outlined in Kotikanyadanam et al.(Kotikanyadanam, Göktepe, and Kuhl 2010). The network was connected to the heart muscle at the end of the network branches, where individual one-dimensional electrical conduction (DC1D2) elements that resemble “resistors” are defined (Bordas et al. 2012).

The electrical response of the tissue is characterized by an action potential, ϕ, and recovery variable, r. The action potential and recovery variable are described by Hurtado and Kuhl (Hurtado and Kuhl 2014).

The global electrical analysis assumes a monodomain response:

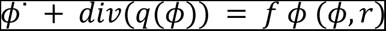

where the flux term, q, characterizes the propagating nature of the electrical waves:

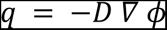

The term D is a second-order diffusion tensor, which can account for anisotropic diffusion. The source term, f *^ϕ^*, characterizes the local action potential profile:

The local biochemical portion of the analysis is modeled through a temporal evolution of the recovery variable, r.

The electrical properties are calibrated to provide physiologically observed activation times. The bundle of His and Purkinje fibers are assigned conduction parameters that generate the physiologically observed wave propagation pattern within the heart whereby the electrical signal first travels down the ventricular septum to the apex and then up the ventricular side walls.

Conduction velocity of the bundle branches were controlled to simulate electrophysiological asynchronies such as LBBB and RBBB (**Figure 2** and Supplemental Video 1) following the literature (Peirlinck et al. 2021). The conduction velocity of the one-dimensional electrical elements of the right bundle branch was reduced while simulating RBBB. Similarly, the conduction velocity of the left bundle branch was reduced simulating LBBB. Tissue responses were assumed to be unchanged.

### 2.2. Structural modeling

Structural modeling of the heart requires complex material models with the consideration of active material response governed by the electrical impulses, and viscoelastic behavior and local fiber orientation through the thickness of the heart wall to be able to simulate realistic cardiac motion. Accordingly, the structural heart was modeled using two types of anisotropic hyperelastic formulation based on that proposed by Holzapfel and Ogden (Holzapfel and Ogden 2009). The passive part of the stress tensor is solely governed by mechanical deformation, while the active part is generated by excitation-induced contraction of myocytes during the course of depolarization. The active stress in the cardiac muscle fiber direction is defined by a time varying elastance model (Walker et al. 2005).

The heart structure has a complex orientation as the fibers that make up the heart are not oriented in the same direction across the surface or through the thickness of the heart wall. To accurately represent the directional properties of the cardiac tissue, an anisotropic material model is employed, which necessitates assigning a local orientation to each element. The fiber angles in the ventricles are defined as approximately –60° for the epicardium and +60° for the endocardium following Streeter et al. (Streeter Jr et al. 1969). Further details can be found in the Simula Living Heart manual. (*SIMULIA Living Heart Human Model User Guide* 2018)

### 2.3. Simulation parameters

The simulation was carried out in two stages (electrical and structural). In the first stage, cardiac conduction was simulated following the steps described in section 2.1. The electrical impulse results obtained from the electrical simulation was used as boundary conditions to simulate the realistic structural heart contraction in Abaqus Explicit 2019 solver (SIMULIA, Dassault Systèmes, Providence, RI). Preexisting LBBB and RBBB conditions (**Figure 3** and Supplemental Video 2) were modeled using the electrical impulse results from the modified electrical simulations as described in section 2.1. The structural simulation was divided into 6 steps including crimping, placement, deployment, preload, beat and recovery (**Figure 4** and Supplemental Video 3). Beat and recovery steps were repeated sequentially for two more cardiac cycles.

The TAVR stent was modeled and meshed using hexahedra-based structured mesh. A superelastic Nitinol alloy (14 constants user material VUMAT available in Abaqus) material model was employed (Morganti et al. 2016; Ghosh et al. 2020a; Oks et al. 2023) for the TAVR stent. Frictional hard contact with a friction coefficient of 0.7 (Rocatello et al. 2018) was used to model the interaction of the TAVR prosthesis with the native tissue. Contact-based interaction was defined between the TAVR prosthesis and the native tissue during the cardiac cycles that allowed the prosthesis to slide and reposition due to the cardiac motion. 1e^-07^ Mass scaling was employed during the deployment stage of each model. During the cardiac cycles, the mass scaling was calibrated to maintain a stable time increment >2.5e^-06^ seconds resulting in an initial added mass of 0.57%, substantially improving performance with minimal impact on results (*SIMULIA Living Heart Human Model User Guide* 2018).

The His bundle (HB) is positioned in between the atrioventricular membranous septum and posterior crest of the muscular septum below the interleaflet triangle of non-coronary leaflet (NCL) and left-coronary leaflet (LCL) (Piazza et al. 2008). Therefore, the area of interest (AOI) was defined below the MS landmark in between NCL and LCL to perform analyses on the stresses and contact-based parameters associated with post-TAVR CCA (**Figure 5**), following the technique described in (Rocatello et al. 2018).

**Figure 5:**
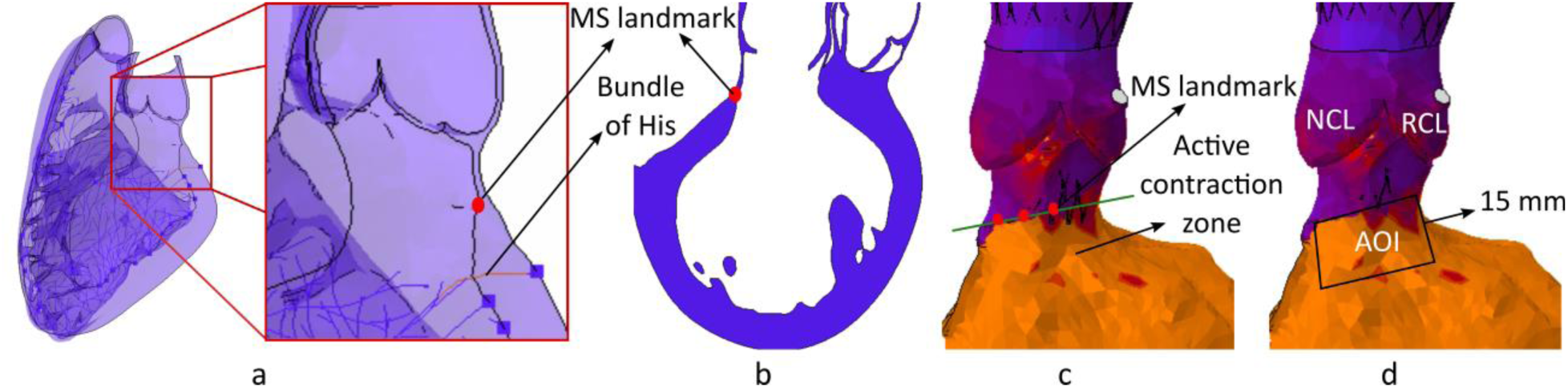
Illustration of the MS landmark from (a) isometric, and (b) long axis view. Identified MS landmark in shown in green (c) and identified area of interest is confined to a black box (d).

## 3. Results

TAVR prostheses were implanted inside the aortic annulus of the living heart model in three preexisting cardiac conduction scenarios (normal cardiac conduction, LBBB and RBBB) and three implantation depths (4mm, 6mm, and 8 mm). Three cardiac cycles were simulated following the TAVR deployment to analyze the interaction between the TAVR prosthesis and AOI. Initially, the stress components (axial, radial, and hoop stress) on the AOI were observed for each patient case and compared with the stress components on the AOI of a healthy heart case which is denoted as ‘No TAVR’. Contact force (**Figure 6**), contact pressure, contact pressure index (CPI) were also analyzed for each virtual patient case.

**Figure 6:**
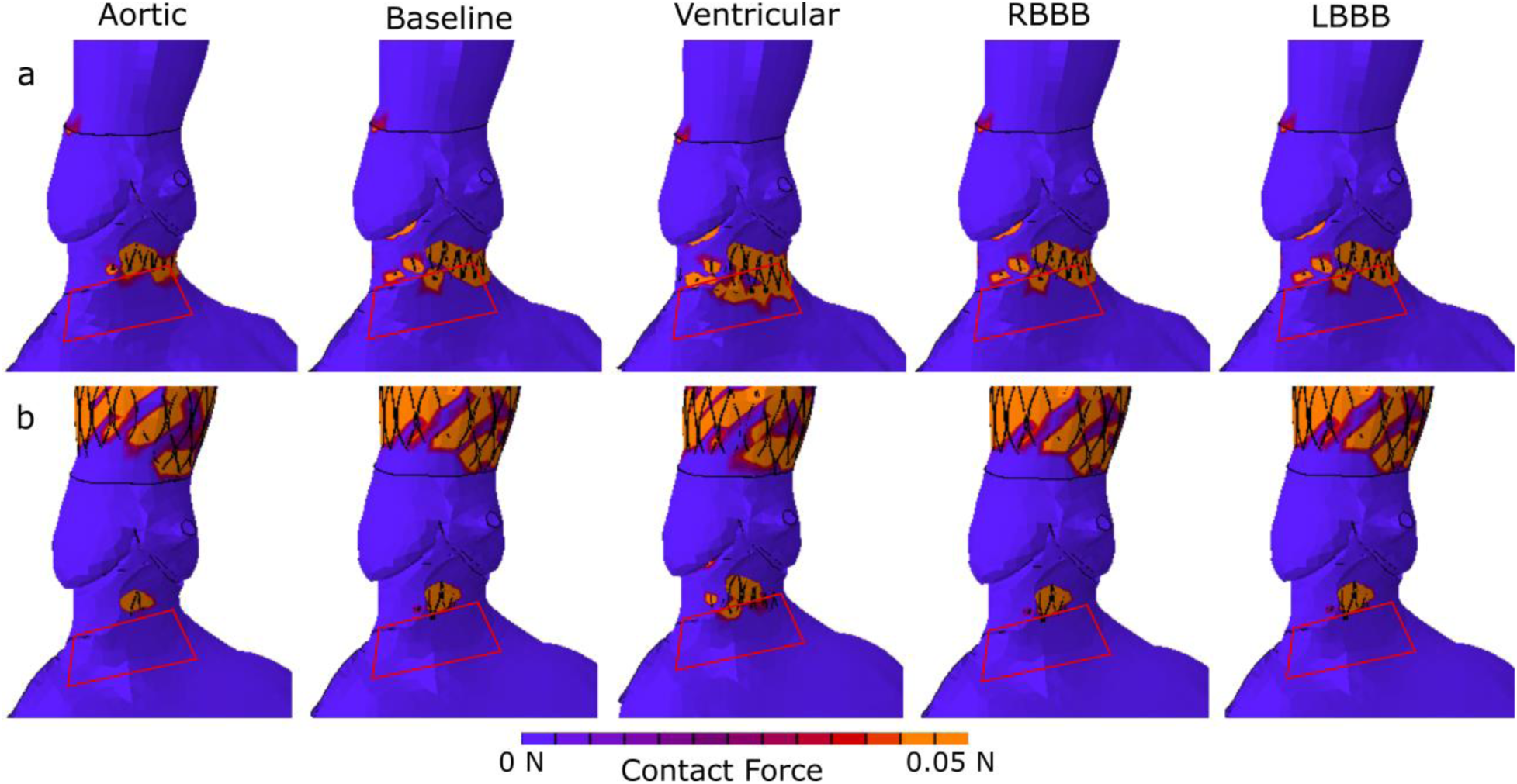
Contact force contour at the AOI during the (a) deployment stage, and (b) late beat stage. The red box depicts the AOI.

### 3.1. Impact of TAVR implantation on the AOI

The principal stress components on the AOI such as axial, radial, and hoop stresses for the ‘No TAVR’ case peak during the late systole of each cardiac cycle. **Figure 7**a shows that TAVR implantation increases all three stress components. The temporal sum of the stress components during peak systole was measured to account for both the magnitude and the duration of the peak (area under the peak stress component). For aortic implantation, there was a 78% increase in the temporal sum of peak axial stress, a 33% increase in radial stress, and a 10% increase in hoop stress. In the baseline implantation depth (ID), the temporal sum of peak axial stress increased by 89%, radial stress by 36%, and hoop stress by 11%. For ventricular implantation, there was a 95% increase in the temporal sum of peak axial stress, a 38% increase in radial stress, and a 13% increase in hoop stress as illustrated in **Figure 7**a.

**Figure 7:**
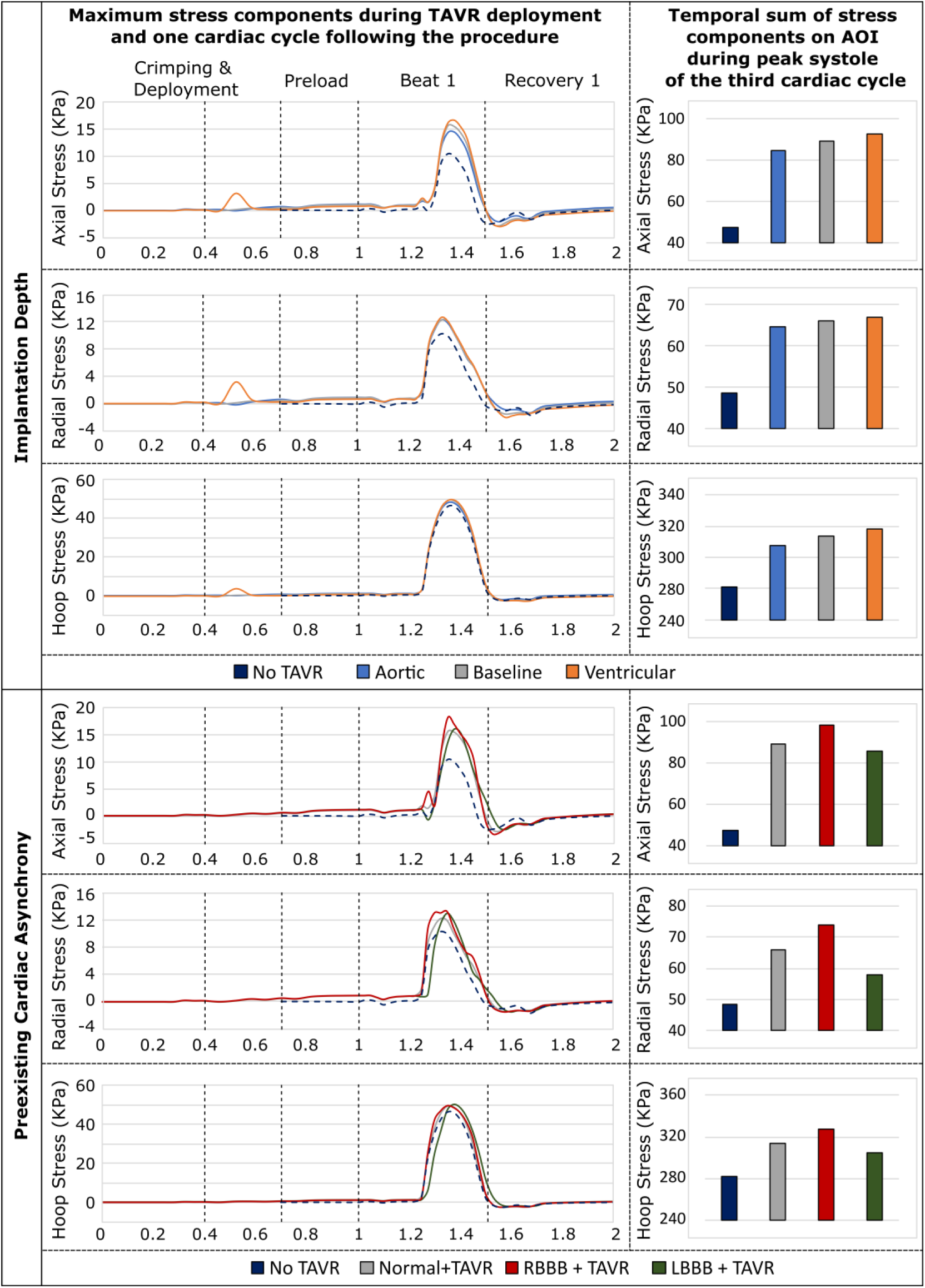
Maximum principal stress components during TAVR deployment and one cardiac cycle following the procedure (left), and the temporal sum of stress components on AOI during peak systole of the third cardiac cycle (right) for (a) three implantation depths, and (b) three cardiac conduction scenarios are compared to the healthy heart.

Stress components on the AOI in preexisting cardiac asynchrony cases (LBBB and RBBB) were also compared to the ‘No TAVR’ case. It was observed that the stress components peaked for a shorter duration in the LBBB case compared to the normal heartbeat case. In the preexisting RBBB case, there was a 10% increase in the temporal sum of peak axial stress, a 12% increase in radial stress, and a 5% increase in hoop stress. In contrast, the preexisting LBBB case showed a relatively smaller increase of 4% in the temporal sum of peak axial stress, 12% in radial stress, and 3% in hoop stress (**Figure 7**b).

Contact-based parameters were also analyzed and compared for all five cases considered in the study. A large peak of maximum contact pressure was observed during the deployment step, which stabilized by the end of the deployment stage and during the preload. However, during the heartbeats following TAVR deployment, there was a fluctuation of contact pressure, with a peak of contact pressure observed from late beat to early recovery step for each cardiac cycle.

### 3.2. Implantation Depth Analysis

The Evolut® 26 mm device was implanted in three different depths (4 mm, 6mm, and 8mm) as described in section 2 Contact pressure, contact force, and contact area index were analyzed for these three implantation depths. The instantaneous contact pressure waveform on the AOI at each simulation step was compared and illustrated in **Figure 8**a. The contact pressure throughout the simulation was significantly lower for the aortic deployment and higher for the ventricular deployment compared to the baseline implantation (**Figure 8**a, b and Supplemental Video 4). The mean of the temporal maximum contact pressure and cumulative contact pressure were measured to be 0.13 KPa, and 17.87 KPa, respectively for aortic deployment, 2.12 KPa, and 298.32 KPa, respectively for baseline implantation, and 3.73 KPa, and 525.78 KPa respectively for ventricular deployment.

**Figure 8:**
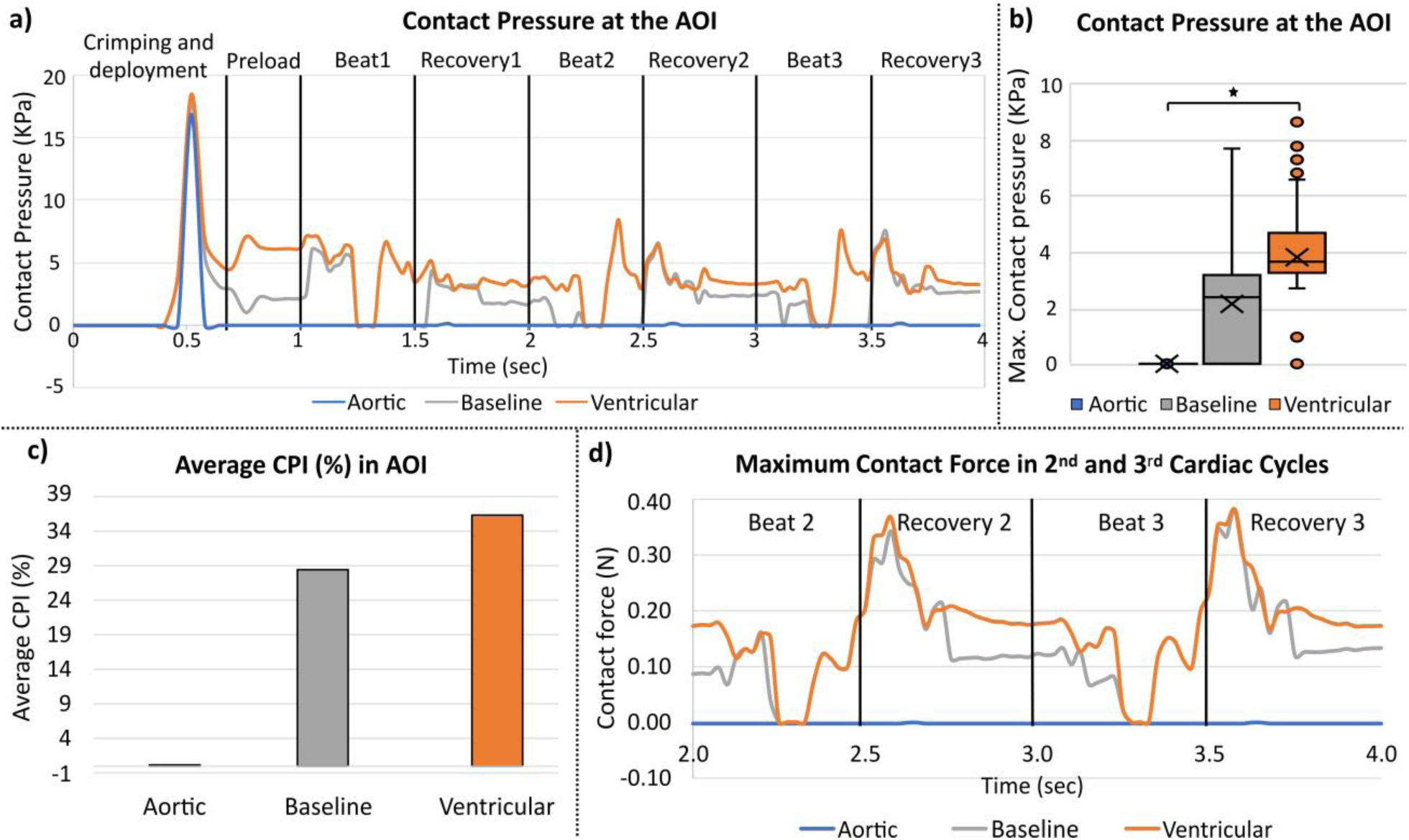
Effects of implantation depth (a) Contact pressure on the AOI at each step, (b) Box plot diagrams of contact pressure at the AOI during three cardiac cycles following TAVR implantation for three implantation depths. Kruskal-wallis test was performed between the three implantation depths yielding P<0.001. (c) Average CPI during three cardiac cycles, and (d) maximum contact force during the 2^nd^ and 3^rd^ cardiac cycles for the three implantation depths.

The temporal average of the contact pressure index (CPI) was measured to be 0.5%, 28%, and 36% for aortic, baseline, and ventricular implantation respectively (**Figure 8**c). Maximum contact force during the cardiac cycles following the TAVR deployment was significantly higher for baseline implantation and ventricular implantation compared to aortic implantation and marginally higher for ventricular implantation compared to baseline implantation. Maximum contact force for aortic, baseline, and ventricular implantation depths were measured as 0.45 N, 2.34 N, and 2.18 N, respectively during the deployment and 0.0003 N, 0.37 N, and 0.38 N, respectively during the cardiac cycles following the TAVR deployment (**Figure 8**d).

### 3.3. Cardiac Asynchrony Analysis

Evolut^®^ 26 mm prosthesis was deployed at a 6 mm implantation depth in preexisting LBBB and RBBB conditions to measure the effect of preexisting cardiac asynchrony on contact-based parameters. The results were then compared to the contact-based parameters measured at a normal heartbeat condition.

The electrical impulse was measured at the free wall of the right ventricle for RBBB case and at the free wall of the left ventricle demonstrating the delay in the conduction of the electrical impulse (**Figure 2**a, b). The distribution of the electrical impulse during one beat is illustrated in **Figure 2**c for Normal heartbeat, LBBB and RBBB cases. The structural changes caused by the preexisting CCAs (RBBB and LBBB) are illustrated from long axis and short axis in **Figure 3**. The qualitative comparison demonstrated a delayed contraction of LV for the LBBB case and a delayed contraction of RV for the RBBB case. Moreover, preexisting LBBB demonstrated a 4.7 mm apex shift towards the left, and preexisting RBBB demonstrated a 3.7 mm apex shift towards the right. The instantaneous contact pressure waveform on the AOI at each simulation step was compared and illustrated in **Figure 9**a. The contact pressure throughout the simulation was higher for the preexisting RBBB and lower for the preexisting LBBB compared to the normal conduction case (**Figure 9**a, b). Preexisting RBBB case has demonstrated a higher mean of the temporal maximum contact pressure, and cumulative contact pressure during 3 cardiac cycles following TAVR (2.4 Kpa, and 339.24 Kpa respectively) compared to the normal heartbeat (2.12 Kpa, and 298.32 Kpa) and preexisting LBBB cases (1.78 Kpa, and 250.27 Kpa, respectively).

**Figure 9:**
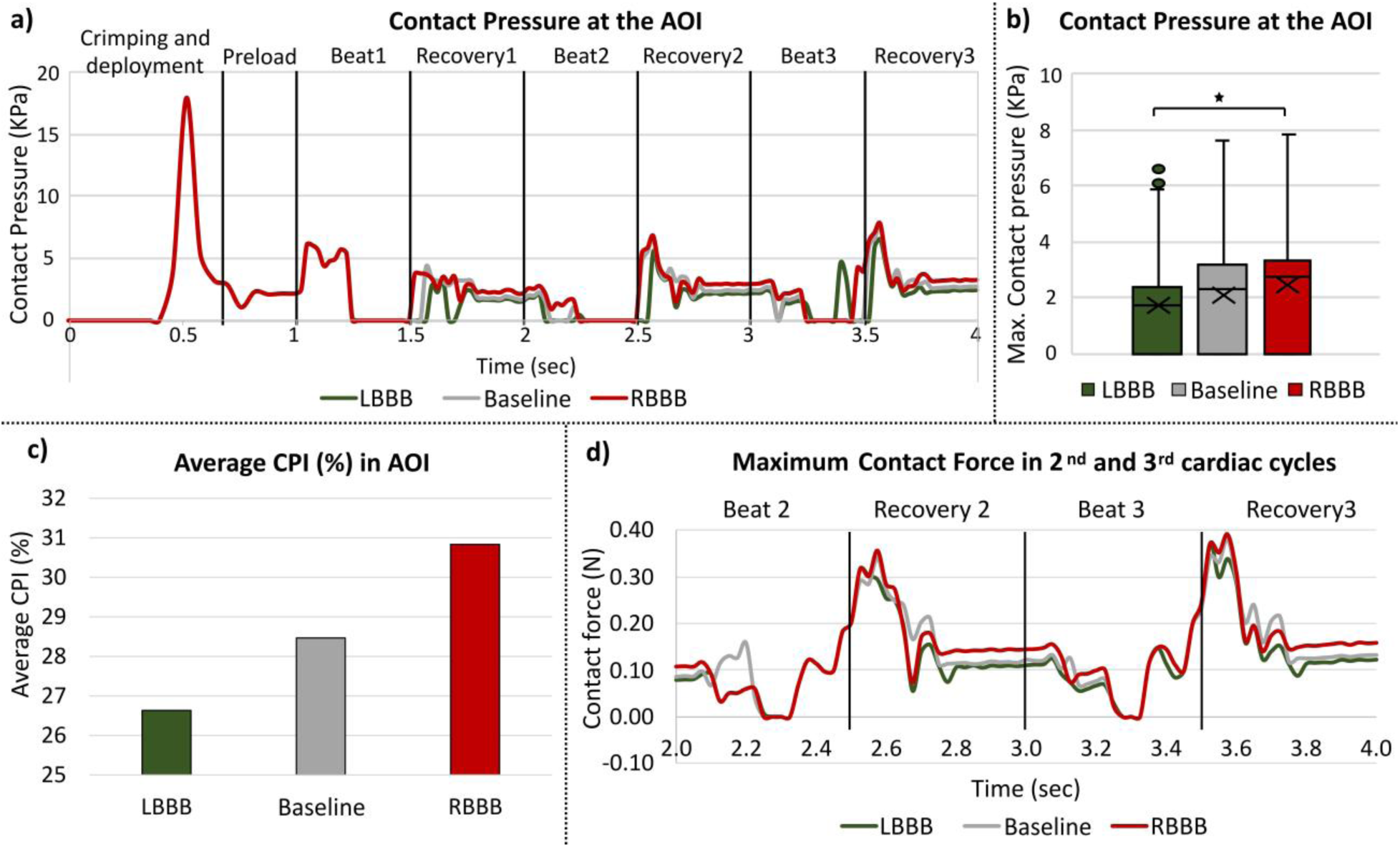
Effects of preexisting Cardiac Asynchrony (a) Contact pressure on the AOI at each step, (b) Box plot diagrams of contact pressure on the AOI during three cardiac cycles following TAVR implantation for three cardiac conduction scenarios. Kruskal-wallis test was performed between the three cardiac conduction scenarios yielding P<0.001. (c) Average CPI during three cardiac cycles, and (d) maximum contact force during the 2^nd^ and 3^rd^ cardiac cycles for the three cardiac conduction scenarios.

The temporary average of the contact pressure index (CPI) was measured to be 26%, 28%, and 30% for LBBB, normal heartbeat, and RBBB cases, respectively (**Figure 9**c). Maximum contact force for LBBB, normal heartbeat, and RBBB cases were unchanged during the deployment stage. However, experienced different contact forces during the cardiac cycles following the TAVR deployment (0.37 N, 0.38 N, and 0.39 N, respectively) as shown in **Figure 9**d.

## 4. Discussion

Post-TAVR cardiac conduction abnormalities (CCA) emerge as a significant post-procedural challenge that may necessitate permanent pacemaker implantation (PPI). The need for PPI has been associated with increased mortality rates and heart failure-related hospitalizations that may hamper the overall success and expansion of TAVR technology. To address the risk of post-TAVR CCA, extensive clinical investigations have been undertaken, delving into various anatomical and procedural variables. Among these factors, implantation depth, Membranous septum (MS) length, ΔMSID, and preexisting RBBB have emerged as prominently linked to post-TAVR CCA. Nevertheless, comprehending the intricate interplay between dynamic heart conditions, preexisting cardiac asynchrony, and the interaction between conduction fibers and the TAVR prosthesis remains a complex challenge. This study introduces a pioneering approach, employing a high-fidelity, electro-mechanically coupled beating heart model to unravel the mechanisms underpinning post-TAVR CCA and their intricate interrelation with anatomical and procedural factors.

The implantation depth holds clinical significance in relation to post-TAVR CCA. This parameter was chosen to elucidate mechanical changes stemming from differing implantation depths and to compare these observations with real-world clinical data. Utilizing the Evolut^®^ 26 mm self-expandable TAVR prosthesis, the research revealed increased stress components within the area of interest (AOI) following TAVR implantation compared to the baseline, healthy condition. Analyzing the contact-based parameters indicated that the peak contact pressure coincides with the deployment phase and stabilizes during successive cardiac cycles. Mean contact pressure at the AOI during each beat and recovery stage showed lower values for aortic deployment and higher values for ventricular deployment compared to baseline implantation. Additionally, maximum contact force and contact pressure index (CPI) exhibited similar trends of implantation depth-dependent variations. Interestingly, stress components and contact-based parameters peaked at distinct phases of the cardiac cycle, underscoring the value of dynamic heart simulations in assessing the risk of post-TAVR CCAs. Synthesizing clinical evidence with prior *in-vivo* and *in-silico* studies, it’s evident that structural dynamics of the heart and contact between the prosthesis and conduction fibers jointly contribute to conduction delays. This study also unveiled a consistent increase in both stress and contact-based parameters with deeper implantation, aligning with clinical observations.

Prior research has established a strong correlation between preexisting RBBB and post-TAVR CCAs. However, the mechanisms underlying this connection remain enigmatic. This study introduces preexisting cardiac asynchrony, encompassing both LBBB and RBBB, as a second parameter. The study investigates the interaction between the TAVR prosthesis and conduction fibers throughout deployment and cardiac cycles, considering the context of cardiac asynchrony. The results revealed delayed conduction and structural shifts due to preexisting LBBB and RBBB. LBBB led to delayed left ventricle (LV) contractions, while RBBB induced delayed contractions in the right ventricle (RV). These structural adaptations, combined with altered electrical impulse conduction, influenced contact-based parameters. Importantly, stress components on the AOI peaks were of shorter duration in the presence of preexisting LBBB compared to a scenario with a normal heartbeat (baseline case). In contrast, preexisting RBBB amplified the cumulative temporal peak stress components on the AOI. These findings underscore the impact of preexisting conduction irregularities on stress distribution following TAVR deployment. The structural alterations associated with RBBB introduce mechanical factors that heighten the risk of post-TAVR CCAs, reinforcing the need to consider preexisting conduction anomalies when assessing these risks.

Evaluating the possibility of early CCA and the likelihood of PPI is of substantial clinical significance. Analysis of the stresses exerted on the AOI clearly indicates that it leads to a distinct surge in contact pressure and force during the TAVR deployment phase. This illustrates how the risk of a new onset of perioperative left bundle branch block (NOP-LBBB) may arise following TAVR. However, an upward shift in the contact area occurred during the successive cardiac cycles, subsequently resulting in nearly zero contact pressure experienced by the AOI which suggests that for this specific patient, a rapid cardiac adaptation occurred that may not require PPI. Baseline and ventricular implantation on the other hand demonstrated a reduced yet steady and cyclic contact pressure during the cardiac cycles. Previously, cyclic stretches on the Purkinje fibers were found to cause conduction delay which indicates that steady and cyclic contact pressure can cause stretches on the conduction fibers leading to conduction delay. This observation may provide insights into the recovery of perioperative CCA and delayed complete AV-block following TAVR. Moreover, by integrating patient-specific electrophysiological conditions and analyzing mechanical parameters during cardiac cycles post-TAVR deployment, it might be feasible to predict which patients will recover from early CCA and thereby evade unnecessary PPI. Such an approach could potentially reduce costs and hospital stays linked to unnecessary interventions.

While static models have been employed for rapid CCA risk assessments and have demonstrated some consistency with clinical findings, they lack the inherent dynamics of a pulsating heart and do not account for preexisting conduction issues. Dynamic models, on the other hand, provide the opportunity for a more physiologically realistic analysis of the emergence of CCAs by considering patient-specific electrophysiological conditions and cardiac cycles. The analysis revealed that the stent frame exerts the highest contact pressure at the early recovery phase of a cardiac cycle. Therefore, reconstructing the patient model at this stage might enable the capture of a snapshot depicting the maximum contact pressure during a cardiac cycle using a static model instead of a dynamic one. This has the potential to improve the assessment of post-TAVR CCA risk, even when utilizing static models.

In summary, dynamic models point out the importance of implantation depth, cardiac asynchrony, and the heart’s dynamics in post-TAVR CCA risk evaluation. The results revealed that the risk of post-TAVR CCA increases with deeper implantation depth, and preexisting RBBB causes increased stresses and contact pressure for a longer period of time enhancing the risk of post-TAVR CCA. Consequently, a cautious approach is recommended when selecting a TAVR device for patients with preexisting RBBB. The presence of preexisting LBBB on the other hand resulted in decreased stresses and contact pressure on the AOI, indicating a reduced likelihood of additional conduction disturbances which is aligned with clinical studies (Maier et al. 2021). The innovative framework presented here has the potential to enhance preprocedural planning and contribute to the development of new TAVR devices, aiming to mitigate the risk of post-TAVR CCA and ultimately enhance patient outcomes.

## 5. Limitations

The Living Heart Model used for electro-mechanical coupling in this study was unidirectional, excluding the potential for mechano-electrical feedback. Nevertheless, a recent study has demonstrated that such exclusion induces only a slight change in conduction velocity (Costabal et al. 2017). This suggests that the one-way electro-mechanical coupling is a reasonable approximation for evaluating post-procedural cardiovascular device risks. This study serves as a methodological approach where a standard Living Heart Model of a healthy heart of a 26-year-old male was used, that can then be manipulated to include various clinical scenarios-such as the TAVR induced CCAs presented in this study, and preexisting conditions such as LBBB and RBBB cases. The use of patient-specific beating heart models is necessary to establish a threshold of the analyzed mechanical parameters that enables to distinguish between patients who may or may not develop a new conduction abnormality following a TAVR procedure. We plan to expand our current simulations to incorporate patient-specific features such as the aortic root and aortic valve calcifications in the future.

## 6. Conclusions

In this study, a novel computational technique is employed to understand the interaction between the TAVR prosthesis and the conduction fibers leading to the assessment of post-TAVR CCA risk in different procedural and electro-mechanical scenarios. The study indicated that a deeper implantation location and a preexisting RBBB causes higher stresses and contact pressure on the conduction fibers and the A-V node, leading to increased risk of post-TAVR CCA. The analysis indicates that implantation above the A-V node MS landmark yielding a negative ΔMSID, reduces the CCA risk and the potential need of post-procedural PPI. Our study also indicates that a conservative approach needs to be considered when choosing a TAVR device for patients with a preexisting RBBB since these patients may experience enhanced stresses and contact pressure due to the dynamic modifications caused by the device deployment and its interaction with the RBBB. This method, employing a systematic digital twin approach, offers potential for personalized preprocedural planning under conditions that simulate a realistically beating heart, aiming to enhance patient outcomes. Additionally, it can be applied to the development of innovative TAVR devices designed to reduce the risk of post-TAVR cardiac conduction abnormalities (CCA).

## Supporting information

Supplemental Video 1(Cardiac Conduction)

Supplemental Video 2 (Structural Adaptation)

Supplemental Video 3 (TAVR Implantation)

Supplemental Video 4 (Contact Force)

## Data Availability

The data is available within the body of the manuscript and the supplemental data

## Acknowledgement

SIMULIA Living Heart Project has provided the base framework of 4-chamber beating heart model employed in this study. The authors thank Dr. Matteo Bianchi for initiating the project and providing scientific insights.

## Acronyms

TAVR: Transcatheter aortic valve replacement
CCA: Cardiac conduction abnormality
MS: Membranous septum
PPI: Permanent pacemaker implantation
A-V: Atrioventricular
AOI: Area of interest
HB: His bundle
CPI: Contact pressure index
CTA: Computed tomography angiography
LBBB: Left bundle branch block
RBBB: Right bundle branch block
RCL: Right coronary leaflet
NCL: Non-coronary leaflet
LCL: Left coronary leaflet

## Statements and Declarations

Author D.B. has an equity interest in Polynova Cardiovascular Inc. Author B.K. is a consultant for Polynova Cardiovascular Inc. The other authors declare that they have no competing interests.

